# Childhood Conduct History is Linked to Amygdalohippocampal Changes in Healthy Adults: A Neuroimaging Behavioral Study

**DOI:** 10.1101/2021.01.20.21250107

**Authors:** AmirHussein Abdolalizadeh, Kamyar Moradi, Mohammad Amin Dabbagh Ohadi, FatemehSadat Mirfazeli, Reza Rajimehr

## Abstract

**Background:** Conduct Disorder (CD) is defined as aggressive, antisocial, and rule-breaking behavior during childhood, and a major risk factor for developing an antisocial personality disorder. However, nearly half the patients develop into seemingly normal status. We aimed to identify psychiatric, emotional, and brain volumetric and functional footprints of childhood CD in healthy young adults with a prior history of CD.

**Methods:** 40 subjects with a prior history of CD (CC) and 1166 control subjects (HC) were identified from the Human Connectome Project. Their psychiatric, emotional, impulsivity, and personality traits were extracted. An emotion task fMRI activation of amygdala and hippocampus, as well as whole-brain and hippocampal/amygdalar segmentation volumetry were analyzed. We then statistically assessed the between-group differences and associations between the assessments and the hippocampal or amygdala nuclei measurements.

**Results:** After correcting for multiple comparisons, we found higher anger aggression, antisocial personality problems, aggressive and rule-breaking behaviors, anxiety, attention-deficit/hyperactivity, intrusive, externalizing, neuroticism, and lower agreeableness in the CC group. The neuroimaging analysis also revealed larger subregions of the left hippocampus in CC group. Significant group × assessment association was found for aggression and left hippocampal presubiculum and basal nuclei of left amygdala.

**Discussion:** Healthy young adults with a prior history of CD still exhibit some forms of antisocial-like behavior, without evidence of emotional recognition disturbances, and with larger left hippocampal subregions. These larger hippocampal and amygdala volumes may play a protective role in CC subjects.

## 1. Introduction

Conduct disorder (CD) is a serious neurodevelopmental disorder defined by a persistent pattern of violence and flouting rules and social norms that significantly impact families and society [1, 2]. CD may accompany many psycho-developmental issues such as substance abuse, depression, anger problems, and lack of empathy, which is the hallmark for callous-unemotional (CU) traits. It is also a major risk factor for developing antisocial personality disorder (ASPD) [2, 3].

Similar patterns of functional or structural disturbances have been seen in both CD and ASPD. For example, functional MRI (fMRI) studies in patients with ASPD suggest significant deficits in the networks related to social cognition and responses to threats [4]. In line with this finding, reduced amygdala response in tasks involving emotions has also been seen in CD [5]. Structural MRI studies also supported a lack of social cognition and emotion in CD subjects. There is evidence of decreased gray matter in various parts of the brain associated with such responses, including lower volumes of left amygdala, right insula, left medial superior frontal gyrus, and left fusiform gyrus in CD patients [3, 6]. On the other hand, some studies observed increased gray matter of the orbitofrontal cortex while the others reported reduced volume comparing CD adults with healthy subjects [7, 8]. Also, voxel-based analysis of patients with ASPD revealed grey matter reduction in the left frontopolar cortex and the paralimbic region associated with memory and emotional problems in these subjects [9, 10].

Structural and functional changes in the orbitofrontal-paralimbic system and hippocampal-amygdala complex both in CD and ASPD suggest them as a possible prognostic neural marker for children with CD for developing into ASPD in the future [11, 12]. However, there is a lack of research regarding neural changes and socio-emotional profiles in healthy young adults with a history of CD in their childhood. In one of the few studies in this regard, women with prior history of CD have reduced grey matter volume in the hippocampus, associated with antisocial behavior [13].

Therefore, we aimed to investigate the psychiatric and socio-emotional aspects of current healthy adults who had a prior history of CD in their childhood, detecting any possible deviation from normal psychiatric or emotional states. Moreover, we aimed to identify neural footprints of childhood CD by evaluating whole-brain volumetric measures, hippocampal subfields, and amygdala nuclei, their activity during an emotional task-fMRI, and their respective associations with psychiatric and emotional evaluations. These measurements may lead us to identify the neural underpinnings of the different neurodevelopmental pathway which encourages the conversion of CD into healthy adults rather than developing ASPD.

## 2. Materials and Methods

### 2.1. Participants

We used the Human Connectome Project young adults (HCP-YA; http://db.humanconnectome.org/) for the current study [14]. It includes 1206 young adults from 25 to 36 years old who have no current psychiatric or neurologic disorders. They have acquired high-resolution structural, diffusion-weighted, and functional MRI data, plus several neuropsychological and cognitive tests and tasks. We identified 40 subjects who had a diagnosis of conduct disorder in their childhood (CC) based on the DSM (The Diagnostic and Statistical Manual of Mental Disorders) conduct disorder criteria extracted from Semi-Structured Assessment for the Genetics of Alcoholism (SSAGA) questionnaires [15] answered by the participants. The antisocial history part of the SSAGA obtained by HCP includes 24 questions regarding the presence of conduct-, or antisocial-like behavioral patterns during childhood and adolescence, their prevalence, duration, and starting date. We identified questions corresponding to the DSM’s conduct disorder criteria and their duration. The subjects scoring the minimum three out of fifteen criteria, with at least one symptom present for more than six months were considered to have conduct disorder history in their childhood (Supplementary Material, Table 1). The remaining subjects who did not have history of conduct disorder in their childhood were considered as the control group (n = 1166; HC).

### 2.2. Evaluations

#### 2.2.1. Demographics

Subject demographics included age, gender, and years of education provided by HCP. We included total household income level as an indicator of socioeconomic status of the subjects, categorized into eight levels from one to eight: 1= less than 10000$, 2 = 10000 to 19999$, 3 = 20000 to 29999$, 4 = 30000 to 39999$, 5 = 40000 to 49999$, 6 = 50000 to 74999$, 7 = 75000 to 99999$, and 8 = 100000$ and more. Penn Progressive Matrices (PMAT) was also included as a measure of fluid intelligence [16].

#### 2.2.2. Emotional

We used the NIH Toolbox Emotion Battery (NIHTB-EB) [17] and Penn Emotion Recognition Task (ER40) [18] for evaluating the emotional aspects of the subjects. The NIHTB-EB includes a variety of tests evaluating negative affect (sadness, fear, anger), psychological well-being (positive affect, life satisfaction, meaning and purpose), social relationships (social support, companionship, social distress, positive social development), and stress and self-efficacy (perceived stress, self-efficacy). In the ER40, each subject is asked about the emotion of a set of 40 faces in five categories: angry, sad, happy, fear, and neutral. The number of correct responses and median response time for the right answers is measured.

#### 2.2.3. Psychiatric and Life Function

We used scores based on Achenbach Adult Self Report (ASR) [19] to evaluate the psychiatric and life function of the subjects. ASR has two components: the syndrome scale and the DSM-oriented scale. We included the anxious/depressed scale, withdrawn, somatic complaints, thought problems, attention problems, aggressive behavior, rule-breaking behavior, intrusiveness, internalizing and externalizing behaviors, and total ASR scores from the syndrome scale. From the DSM-oriented scale, we included DSM depressive, anxiety, somatic, avoidant personality, attention-deficit/hyperactivity, and antisocial personality problems. We included all the ASR-based scores of the subjects in the current study as gender and age-adjusted T-scores.

#### 2.2.4. Personality

Personality traits of subjects were acquired based on NEO five-factor inventory (NEO-FFI) [20]. This 60-item questionnaire can be used to study the personality construct of an individual to “agreeableness,” “openness,” “neuroticism,” “extraversion,” and “conscientiousness”.

#### 2.2.5. Impulsivity

We used delay discounting task as a measure of impulsivity [21]. In this task, the subjects are given the choice of a reward now or a more valuable reward with a delay. The trials are repeated with changes following a set of rules until an *indifference point* is achieved: the subjective value of the later more valuable reward is equal to the present less valuable reward. Plotting the indifference point reward values against delay time (both normalized) results in a hyperbolic graph. The area under the curve (AUC) of this graph is considered as a measure of impulsivity: higher impulsivity results in smaller present reward values preferred (a.k.a., considered being equal) to a shorter delay interval, resulting in a steeper hyperbolic chart and lesser AUC [22]. We included delay discounting AUCs of trials with 200$ and 40000$ from HCP.

### 2.3. Imaging

#### 2.3.1. Acquisition and Preprocessing of Structural Data

High-resolution T1 and T2 scans were acquired using 3T Connectome Siemens Skyra with the following parameters: T1: 3D-MPRAGE, TR = 2400ms, TE = 2.14ms, TI = 1000ms, FOV = 224×224, Voxelsize = 0.7mm isotropic; T2: T2-SPACE, TR = 3200ms, TE = 565 ms, FOV = 224×224, Voxelsize = 0.7mm isotropic. We used the minimally preprocessed structural data, available as structural extended preprocessed [23]. The analysis pipeline, which resulted in the minimal preprocessed structural data, is briefly as follows: All structural acquisitions were first corrected for gradient distortion, aligned and averaged, skull stripped, corrected for readout distortion, and then fed to the *recon-all* pipeline of FreeSurfer [24], using both high-resolution T1 and T2 weighted images for better identification of pial surfaces. Finally, morphometric measures of cortical parcellations and subcortical segmentations were calculated based on the Deskian-Killiany atlas [25]. The preprocessed data were available for 35 CC subjects. We matched these subjects with 69 HCs based on age, gender, and education using *matchIt* library in R [26].

#### 2.3.2. Hippocampal/Amygdala Subnuclei Segmentation

We used segmentation of hippocampal subfields and nuclei of the amygdala implemented in FreeSurfer v7.1.1 [27, 28]. This automated approach provides the hippocampus and amygdala segmentation based on a high-resolution probabilistic atlas derived from fifteen autopsy samples. We used high-resolution (starting with hires_) T1 and T2 scans provided in the FreeSurfer output */mri* folder in structural extended preprocessed data by renaming the associated files as an input for *segmentHA_T2*.*sh* command (Figure 1). The analysis was run on the 35 CC and 69 HC subjects using the *parallel* command in Linux Ubuntu 18.04 to use all the CPU threads [29]. Divisions of segments were summed to include that segment as a single volume (e.g., CA1 equals CA1 head plus CA1 body).

**Figure 1.**
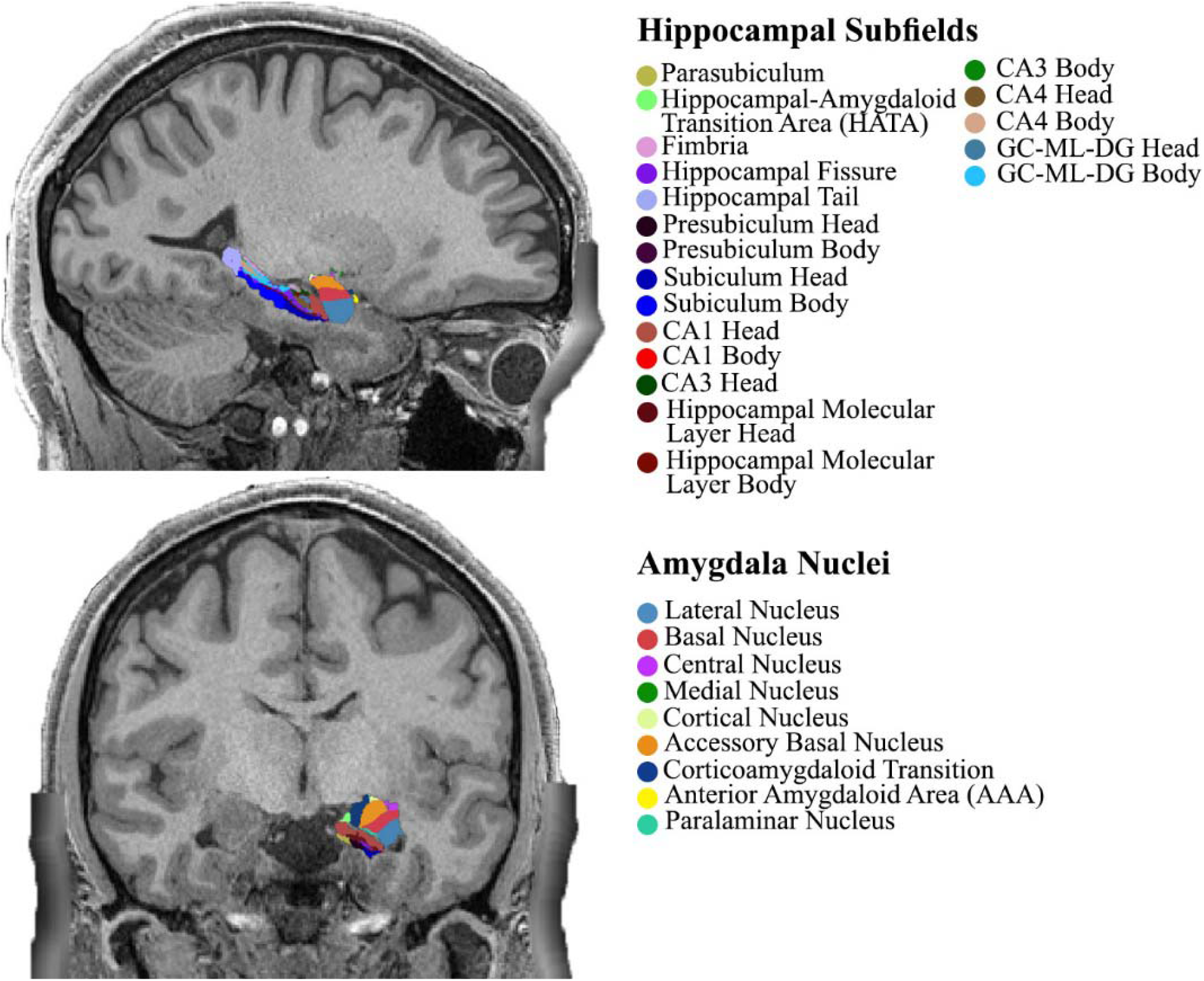
Hippocampal subfields and amygdala nuclei overlaid on sagittal (top) and coronal (bottom) high-resolution T1 scan of a CC subject using *segmentHA_T2*.*sh* script. Different areas are color-coded to be better identified. CA = Cornu Ammonis, GC-ML-DG = Granule Cell and Molecular Layer of Dentate Gyrus.

#### 2.3.3. Emotion Processing Task fMRI

The whole-brain EPI acquisition done by the same 3T Siemens Skyra scanner parameters were as follows: 32 channel head coil, TR = 720ms, TE = 33.1ms, flip angle = 52deg, BW = 2290Hz/Px, FOV = 208×180mm, 2.0mm isotropic voxelsize, with a multi-band acceleration factor of 8. Two task fMRI runs were done, one with right-to-left and another with left-to-right phase encoding directions. The procedure of task fMRI is described elsewhere [30]. The emotion task fMRI is adapted from Hariri et al. [31], in which the subjects were asked to match the two shown pictures of faces or shapes on the lower part of the screen to the one presented above. The faces had either fearful or angry expressions and the shapes were depicting fearful or threatening situations. It was shown by Hariri et al., that face vs. shapes contrast shows a reliably strong activation of the bilateral amygdala indicating selective activation of amygdala to negative facial stimuli. Task fMRI data was minimally preprocessed and made available by HCP [23]. In brief, the data were gradient distortion corrected, motion-corrected, EPI distortion corrected, and then registered non-linearly with the “surface-based” method to MNI registered subject’s T1 scans. The minimally preprocessed data was then fed to FSL FEAT to apply contrasts and derive betas [32]. We used the faces vs. shapes contrast grayordinates stats (cope3.feat/cope1.dtseries.nii) analyzed 2mm FWMH smoothed task fMRI data of 1039 subjects (36 CC and 1003 HC) provided by HCP. Mean beta values of bilateral amygdala and hippocampi were extracted using cifti-matlab commands (https://github.com/Washington-University/cifti-matlab). CIFTI-indices associated for each region is as follows: left amygdala = 59688 to 60002, right amygdala = 60003 to 60334, left hippocampus = 84561 to 85324, and right hippocampus = 85325 to 86119.

### 2.3. Statistical Analysis

We used Rstudio software based on R statistical package version 3.6.1 [33, 34]. Subject demographics and evaluations were compared between 40 CC and the rest of the HCP subjects (HC; n = 1166), using chi-square, t-test, or Mann-Whitney test based on the type and distribution of the data.

To evaluate structural volumetric differences between two groups, FreeSurfer generated cortical parcellation thickness provided by HCP were included in a general linear model (GLM) covarying age and gender. In another GLM, differences between subcortical volumes were evaluated and were adjusted for age, gender, and total intracranial volume. The same model was also applied for hippocampal and amygdala segmentation volumes with the addition of education and subject income level to the covariates. Moreover, we used a GLM evaluating the association between all the evaluations in part 2.2 and the interaction between hippocampal and amygdala segmentation volumes and group variable (hippocampal or amygdala × group) with the same covariates. To overcome the multiple testing problem, we used the Benjamini-Hochberg False Discovery Rate (FDR) method to correct the *p*-values wherever applicable [35]. Corrected *p*-values equal to or less than 0.05 were considered to be statistically significant.

## 3. Results

### 3.1. Demographics

The demographics and their differences are shown in table 1. There were no differences between age, income level, or fluid intelligence between the two groups. However, CC subjects had slightly lower education (mean (SD): 14.30 (1.86) vs. 14.88 (1.82), *p* = 0.046), and higher males vs. controls (M (%): 29 (72.5%) vs. 521 (44.7%), *p* = 0.001).

**Table 1.** Demographics, emotional, and psychiatric evaluations between 40 subjects with prior history of conduct disorder in childhood, and 1166 subjects without history of conduct.

| Variables | HC<br>n = 1166 | CC<br>n = 40 | *p* | Corrected<br>*p* |
| --- | --- | --- | --- | --- |
| **Demographics** |  |  |  |  |
| Age (mean (SD)) | 28.84 (3.70) | 28.80 (3.38) | 0.948 |  |
| Gender (M (%)) | 521 (44.7) | 29 (72.5) | 0.001* |  |
| Years of Education (mean (SD)) | 14.88 (1.82) | 14.30 (1.86) | 0.046* |  |
| Income Level (mean (SD)) | 5.03 (2.17) | 4.38 (2.31) | 0.063 |  |
| PMAT Correct Responses (mean (SD)) | 16.68 (4.92) | 15.97 (4.60) | 0.373 |  |
| **Emotion** |  |  |  |  |
| **Penn Emotion Recognition Test** |  |  |  |  |
| Number of Correct Responses (mean (SD)) | 35.54 (2.56) | 34.85 (3.21) | 0.099 | 0.523 |
| Correct Responses Median Response Time (ms) (mean (SD)) | 1827.45 (326.16) | 1897.47 (312.05) | 0.182 | 0.408 |
| Number of Correct Anger Identifications (mean (SD)) | 6.75 (1.03) | 7.10 (0.84) | 0.034 | 0.209 |
| Number of Correct Fear Identifications (mean (SD)) | 6.90 (1.20) | 6.70 (1.45) | 0.314 | 0.821 |
| Number of Correct Happy Identifications (mean (SD)) | 7.96 (0.23) | 7.97 (0.16) | 0.616 | 0.821 |
| Number of Correct Neutral Identifications (mean (SD)) | 7.13 (1.25) | 6.53 (1.71) | 0.003 | 0.076 |
| Number of Correct Sad Identifications (mean (SD)) | 6.80 (1.14) | 6.55 (1.40) | 0.18 | 0.559 |
| **NIH Toolbox: Emotion Battery** |  |  |  |  |
| Anger Affect (mean (SD)) | 47.90 (8.28) | 51.80 (10.92) | 0.004 | 0.076 |
| Anger Aggression (mean (SD)) | 51.88 (8.69) | 60.47 (10.48) | <0.001 | <0.001* |
| Anger Hostility (mean (SD)) | 50.65 (8.61) | 51.89 (10.52) | 0.374 | 0.692 |
| Emotional Support (mean (SD)) | 51.32 (9.57) | 49.86 (12.04) | 0.347 | 0.843 |
| Fear Affect (mean (SD)) | 50.22 (7.97) | 51.89 (10.92) | 0.199 | 0.527 |
| Fear Somatic Symptoms (mean (SD)) | 52.02 (8.27) | 52.26 (9.63) | 0.859 | 0.884 |
| Friendship (mean (SD)) | 50.20 (9.09) | 50.25 (9.97) | 0.975 | 0.843 |
| Instrumental Support (mean (SD)) | 48.04 (8.95) | 47.49 (12.29) | 0.71 | 0.912 |
| Life Satisfaction (mean (SD)) | 54.41 (9.19) | 53.21 (10.05) | 0.422 | 0.821 |
| Loneliness (mean (SD)) | 51.15 (8.77) | 52.97 (9.30) | 0.198 | 0.466 |
| Meaning and Purpose (mean (SD)) | 51.69 (8.66) | 50.63 (9.38) | 0.448 | 0.692 |
| Perceived Hostility (mean (SD)) | 48.72 (8.65) | 50.72 (9.67) | 0.154 | 0.527 |
| Perceived Rejection (mean (SD)) | 48.68 (8.87) | 50.42 (9.67) | 0.223 | 0.466 |
| Perceived Stress (mean (SD)) | 48.40 (9.11) | 51.00 (10.45) | 0.078 | 0.349 |
| Positive Affect (mean (SD)) | 50.01 (7.83) | 48.92 (9.23) | 0.388 | 0.559 |
| Sadness (mean (SD)) | 46.41 (7.97) | 48.67 (8.87) | 0.08 | 0.362 |
| Self-efficacy (mean (SD)) | 50.80 (8.35) | 51.61 (9.54) | 0.548 | 0.762 |
| **Psychiatry and Life Function** |  |  |  |  |
| **DSM-Oriented Adult Self-Report** |  |  |  |  |
| Depressive Problems (mean (SD)) | 53.97 (5.71) | 56.23 (8.33) | 0.016 | 0.136 |
| Antisocial Personality Problems (mean (SD)) | 52.94 (4.53) | 57.73 (9.79) | <0.001 | <0.001* |
| Anxiety Problems (mean (SD)) | 53.30 (5.19) | 55.52 (7.04) | 0.009 | 0.046* |
| Somatic Problems (mean (SD)) | 54.07 (5.80) | 56.02 (8.85) | 0.04 | 0.164 |
| Avoidant Personality Problems (mean (SD)) | 54.40 (6.08) | 56.25 (8.13) | 0.062 | 0.057 |
| Attention-Deficit/Hyperactivity Problems (mean (SD)) | 54.79 (5.62) | 58.25 (9.40) | <0.001 | 0.021* |
| **Syndrome Scale of Adult Self-Report** |  |  |  |  |
| Aggressive Behavior (mean (SD)) | 52.60 (3.90) | 56.23 (9.31) | <0.001 | 0.007* |
| Rule Breaking Behavior (mean (SD)) | 53.80 (5.01) | 58.62 (9.36) | <0.001 | <0.001* |
| Intrusive (mean (SD)) | 53.75 (5.44) | 56.98 (7.40) | <0.001 | 0.006* |
| ASR Total Score (mean (SD)) | 47.90 (8.84) | 53.05 (12.04) | <0.001 | 0.008* |
| Anxious/Depressed (mean (SD)) | 53.97 (6.24) | 55.95 (8.99) | 0.052 | 0.057 |
| Withdrawn (mean (SD)) | 53.65 (5.75) | 56.38 (9.62) | 0.004 | 0.028* |
| Somatic Complaints (mean (SD)) | 53.96 (5.91) | 56.35 (9.21) | 0.014 | 0.148 |
| Thought Problems (mean (SD)) | 53.70 (5.68) | 56.17 (7.67) | 0.008 | 0.021* |
| Attention Problems (mean (SD)) | 54.90 (5.39) | 57.40 (8.22) | 0.005 | 0.052 |
| Internalizing (mean (SD)) | 48.60 (10.63) | 52.30 (13.71) | 0.032 | 0.136 |
| Externalizing (mean (SD)) | 48.70 (8.76) | 55.85 (12.20) | <0.001 | <0.001* |

Significant differences marked with asterisks. HC = Healthy Controls with no History of Conduct, CC = Childhood Conduct History, M = Male, PMAT: Penn Matrix Test of Fluid Intelligence, DSM = The Diagnostic and Statistical Manual of Mental Disorders, ASR = Adult Self-Report.

### 3.2. Evaluations

#### 3.2.1. Emotion

CC subjects had higher anger aggression (from NIHTB-EB) compared to HC (mean (SD): 60.47 (10.48) vs. 51.88 (8.69), respectively; corrected *p* < 0.001). This measure evaluates aggression as the behavioral component of anger. There were no other significant differences in other components of NIHTB-EB or ER40 between the two groups (Table 1).

#### 3.2.2. Psychiatric and Life Function

CC subjects had significantly greater antisocial personality problems and have reported higher aggressive and rule-breaking behaviors than HCs based on the age and gender-adjusted T-scores of those measures. Moreover, CC subjects had higher anxiety, attention-deficit/hyperactivity problems, externalizing, intrusive and withdrawn behavior, thought problems and ASR total T-scores as well. There were no other significant differences noted between the two groups (Table 1).

#### 3.2.3. Personality

Subjects in CC group had lower agreeableness compared to HC (mean (SD): 30.00 (6.46) vs. 33.44 (5.77), *p* < 0.001). They also had higher neuroticism compared to HC (mean (SD): 19.15 (8.37) vs. 16.72 (7.34), *p* = 0.041). There were no significant differences in other NEO-FFI measurements between two groups, including extraversion (HC vs. CC; mean (SD): 30.64 (5.96) vs. 30.75 (6.26); *p* = 0.908), openness (HC vs. CC; mean (SD): 28.16 (6.27) vs. 29.45 (6.28); *p* = 0.201), and conscientiousness (HC vs. CC; mean (SD): 34.47 (5.91) vs. 33.58 (6.58); *p* = 0.348).

#### 3.2.4. Impulsivity

There were no significant differences between the two groups in Area Under the Curve of delay discounting trials of 200$ (HC vs. CC; mean (SD): 0.26 (0.20) vs. 0.24 (0.23), *p* = 0.608) and 40K$ (HC vs. CC; mean (SD): 0.51 (0.29) vs. 0.45 (0.28), *p* = 0.227).

### 3.3. Imaging

#### 3.3.1. Whole-Brain Volumetric Study

Using morphometric measures generated by FreeSurfer and provided by HCP, there were no differences between the thickness of cortical parcellations based on the Desikan-Killiany atlas (Supplementary Material, Table 2). The statistical model was applied on volumetric measures of FreeSurfer output with the addition of Total Intracranial volume as a covariate. There were no significant differences between CC and HC after Benjamini-Hochberg correction (Supplementary Material, Table 3).

#### 3.3.2. Hippocampal Subfields and Amygdala Nuclei

CC subjects had larger CA3, CA4, presubiculum, GC-ML (Granule cell and molecular layer) of the dentate gyrus, and hippocampus-amygdala transition area (HATA) in the left side compared to the controls using a statistical model adjusted for age, gender, education, income, and total intracranial volume (Table 2). There were no differences between amygdala nuclei or right-side hippocampal subfields between two groups.

**Table 2.** Volumes of hippocampal subfields and amygdala nuclei as cubic millimeters (mean (SD)).

| Volume | HC<br>(n = 69) | CC<br>(n = 35) | *p* | Corrected *p* |
| --- | --- | --- | --- | --- |
| **Amygdala** |  |  |  |  |
| left Lateral nucleus | 654.38 (79.92) | 695.63 (77.06) | 0.121 | 0.266 |
| left Basal nucleus | 451.92 (56.93) | 479.26 (44.49) | 0.064 | 0.186 |
| left Accessory Basal nucleus | 274.31 (35.27) | 290.93 (28.16) | 0.119 | 0.266 |
| left Anterior amygdaloid area | 58.02 (8.16) | 61.33 (7.05) | 0.151 | 0.316 |
| left Central nucleus | 57.09 (9.60) | 58.36 (9.85) | 0.842 | 0.882 |
| left Medial nucleus | 29.84 (6.31) | 30.95 (7.19) | 0.709 | 0.821 |
| left Cortical nucleus | 31.55 (4.63) | 32.87 (4.71) | 0.939 | 0.939 |
| left Corticoamygdaloid transition | 196.91 (26.28) | 211.81 (23.60) | 0.034 | 0.141 |
| left Paralaminar nucleus | 51.58 (6.37) | 54.94 (5.91) | 0.024 | 0.141 |
| left Whole amygdala | 1805.60 (214.27) | 1916.08 (178.72) | 0.062 | 0.186 |
| right Lateral nucleus | 673.14 (78.52) | 721.58 (77.84) | 0.018 | 0.133 |
| right Basal nucleus | 455.62 (57.73) | 476.98 (47.74) | 0.528 | 0.726 |
| right Accessory Basal nucleus | 269.92 (38.06) | 282.16 (28.80) | 0.878 | 0.898 |
| right Anterior amygdaloid area | 62.47 (8.71) | 65.95 (8.26) | 0.338 | 0.533 |
| right Central nucleus | 61.28 (10.83) | 62.78 (11.27) | 0.773 | 0.830 |
| right Medial nucleus | 29.35 (6.60) | 30.44 (5.92) | 0.598 | 0.752 |
| right Cortical nucleus | 28.84 (6.27) | 30.93 (4.23) | 0.640 | 0.782 |
| right Corticoamygdaloid transition | 187.75 (25.18) | 199.46 (22.17) | 0.339 | 0.533 |
| right Paralaminar nucleus | 50.23 (6.16) | 53.04 (5.92) | 0.228 | 0.437 |
| right Whole amygdala | 1818.59 (216.42) | 1923.30 (186.45) | 0.167 | 0.335 |
| **Hippocampus** |  |  |  |  |
| left CA1 | 677.75 (90.02) | 718.59 (89.78) | 0.083 | 0.214 |
| left CA3 | 207.35 (26.44) | 227.47 (32.27) | 0.002 | 0.036* |
| left CA4 | 248.21 (29.40) | 267.58 (29.03) | 0.003 | 0.036* |
| left fimbria | 97.26 (20.65) | 103.53 (20.18) | 0.304 | 0.533 |
| left GC-ML-DG | 302.63 (36.58) | 326.36 (36.47) | 0.003 | 0.036* |
| left HATA | 61.52 (9.23) | 68.38 (10.07) | 0.003 | 0.036* |
| left hippocampal fissure | 133.03 (24.22) | 140.13 (22.34) | 0.328 | 0.533 |
| left Hippocampal tail | 578.67 (62.03) | 611.16 (61.08) | 0.035 | 0.141 |
| left molecular layer | 408.44 (55.31) | 422.29 (54.23) | 0.435 | 0.627 |
| left parasubiculum | 68.15 (11.67) | 74.05 (15.21) | 0.049 | 0.180 |
| left presubiculum | 304.57 (38.69) | 331.69 (39.92) | 0.005 | 0.045* |
| left subiculum | 455.10 (61.79) | 480.28 (53.12) | 0.079 | 0.214 |
| right CA1 | 716.69 (100.82) | 750.26 (96.82) | 0.239 | 0.438 |
| right CA3 | 228.68 (30.78) | 242.54 (33.41) | 0.055 | 0.186 |
| right CA4 | 260.13 (31.77) | 275.26 (30.09) | 0.035 | 0.141 |
| right fimbria | 90.41 (21.01) | 94.73 (18.93) | 0.737 | 0.821 |
| right GC-ML-DG | 313.32 (37.78) | 332.04 (35.20) | 0.030 | 0.141 |
| right HATA | 61.70 (9.40) | 66.39 (11.05) | 0.119 | 0.266 |
| right hippocampal fissure | 135.72 (35.29) | 137.23 (22.37) | 0.746 | 0.821 |
| right Hippocampal tail | 588.62 (62.44) | 610.80 (63.35) | 0.353 | 0.536 |
| right molecular layer | 415.11 (55.42) | 419.86 (53.28) | 0.712 | 0.821 |
| right parasubiculum | 69.22 (13.14) | 73.48 (16.30) | 0.442 | 0.627 |
| right presubiculum | 290.36 (36.59) | 302.17 (41.81) | 0.597 | 0.752 |
| right subiculum | 441.16 (52.51) | 456.46 (51.63) | 0.554 | 0.738 |

*p*-values are for a general linear model to identify between-group differences with age, gender, income level, years of education, and total intracranial volumes as covariates. Corrected *p*-values are corrected using Benjamini-Hochberg method. CC = Childhood Conduct history, HC = No Childhood Conduct History, CA = Cornu Ammonis, GC-ML-DG = Granule Cell and Molecular Layer of Dentate Gyrus, HATA = Hippocampus-Amygdala Transition Area.

#### 3.3.3. Emotion Task fMRI of Hippocampus and Amygdala

Faces vs. shapes contrast beta values of emotion task fMRI was available for 1039 subjects (36 CC and 1003 HC). Using an ANCOVA model and adjusting for age, gender, education, and income level, there were no significant differences between mean beta values of the two groups in the left (HC vs. CC (mean (SD)): 30.22 (18.12) vs. 32.77 (18.60), F (1, 1026) = 0.64, *p* = 0.42) and right (HC vs. CC (mean (SD)): 31.55 (18.57) vs. 29.73 (17.21), F (1, 1026) = 0.27, *p* = 0.60) amygdala. There were also no significant differences between mean beta values of the two groups in the left (HC vs. CC (mean (SD)): 8.69 (14.67) vs. 11.12 (14.07), F (1, 1026) = 0.68, *p* = 0.40) and right (HC vs. CC (mean (SD)): 9.95 (13.04) vs. 10.69 (14.46), F (1, 1026) = 0.09, *p* = 0.75) hippocampus.

#### 3.3.4. Association of Evaluations with Hippocampal Subfields and Amygdala Nuclei Volumes

There was two significant group × volume associations with neuropsychiatric evaluations after correcting all the *p*-values of multiple comparisons (n = 2112) using Benjamini-Hochberg method: ASR aggression age-,gender-adjusted T-score and left basal nucleus of amygdala (Estimate (Std. Error) = −0.10 (0.02), corrected *p* = 0.04) and left presubiculum (Estimate (Std. Error) = −0.14 (0.03), corrected *p* = 0.01). The results are depicted in figure 2. No other significant associations were found.

**Figure 2.**
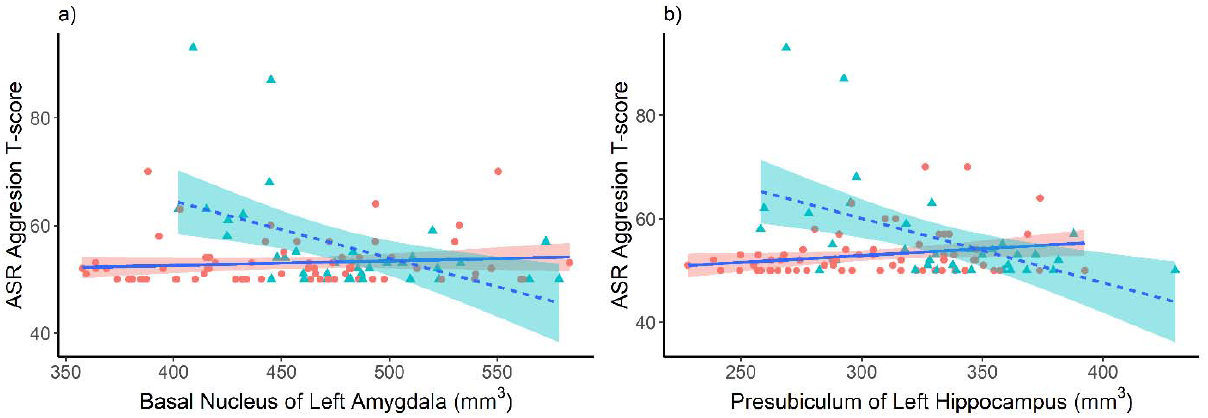
Significant group × assessment associations between Aggression T-scores and basal nucleus of left amygdala (a) and presubiculum of left hippocampus (b) for 35 subjects with prior history of conduct disorder (CC group; Triangles, Green) and 69 subjects with no prior history of conduct disorder (HC group; Circles, Red). Note the negative association between aggression and volumes for CC group. ASR: Adult Self-Report

## Discussion

In the present study, we provided novel evidence regarding the effect of the previous history of CD in childhood on behavioral, emotional, and neurobiological features of currently healthy adults. The behavioral parameters were systematically compared between CC and HC groups in four major domains: emotion, psychiatric and life functions, impulsivity, and personality. Significant differences were reported as higher anger aggression, antisocial personality problems, aggressive and rule-breaking behaviors, anxiety, attention-deficit/hyperactivity, intrusive, externalizing, ASR total T-scores, neuroticism, and lower agreeableness in the CC group. From a neuroimaging perspective, the cortical thickness and volume of none of the brain regions or amygdala nuclei were different between the two groups. However, CC group had higher CA3, CA4, presubiculum, GC-ML, and HATA in the left hippocampus. Interestingly, higher adult self-report aggression T-scores were negatively associated with volumes of the basal nucleus of left amygdala and the presubiculum of left hippocampus in the CC group using a group × volume interaction model. The differences between the groups cannot be attributed to the demographic characteristics since repeated comparisons showed similar features, with both groups sharing a similar demographic backgrounds.

There is robust evidence regarding the significant association between child- and adolescent-onset CD and adulthood antisocial personality disorder [36]. However, childhood CD does not necessarily progress to antisocial disorder as a proportion of subjects with CD experience recovery, especially those with low psychopathic traits [37]. The improvement from CD to the normal condition has been attributed to various biological and environmental factors such as less initial severity of CD, higher child verbal intelligence, greater family socioeconomic status, and not having antisocial biological parents [38]. While this is probable for children with CD to improve to nearly normal and healthy status in case of receiving appropriate cares and treatments, surprisingly, to the best of our knowledge, few studies have ever investigated the behavioral and neurobiological outcomes of this population in adulthood, and this study is considered among the first of its kind in this topic. Our findings suggest that although subjects with CD-to-normal status reversion do not meet the criteria for adult-onset antisocial personality disorder, they still show some antisocial traits. Most of these traits are also evident in antisocial personality disorder (e.g., rule-breaking, aggressive behavior), and the mere difference might be in the trait of the symptoms.

Callous-unemotional (CU) traits are a personality-related subgroup of psychopathic personality traits, characterized by low empathy, restricted affect, interpersonal callousness, and lack of motivation and are significantly associated with more severe and more persistent antisocial behaviors [39]. Although subjects with the previous CD still exhibited some antisocial traits, the emotion and personality domains were found within an almost normal range, confirming the low levels of CU traits in our participants which help their improvement over time in antisocial behaviors.

Literature has addressed alterations in brain structure or structural connectivity of youths with CD, oppositional defiant disorder (ODD), and conduct problems [40]. Particularly, according to a meta-analysis of six studies, youths with child-onset conduct problems showed decreased grey matter volume in the left amygdala and anterior insula compared to typically developing ones [6]. Notably, all previous studies have investigated the alterations in brain structures in patients with current conduct problems. For the first time, this study showed that adults with prior CC and current healthy status exhibit larger regions in the left hippocampus compared to individuals with no history of conduct problems during their lifetime. It was suggested that children with CD have smaller hippocampus compared to matched healthy controls, which was found associated with higher impulsive behaviors [41]. On the other hand, our CC group consists of participants with a history of CD that have experienced behavioral recovery. The larger hippocampus in the CC group might be justified by the hypothesis that the larger hippocampus has a protective role in CD and increases the chance of recovery. The protective role of the hippocampus volume has been mentioned in a number of previous studies [42, 43]. Moreover, we showed that in addition to the enlarged left hippocampus subregions in the normal praticipants with CC, there is a negative association between presubiculum volume and aggressive behaviors. The behavioral correlates of presubiculum has not been studied, however, recent studies regarding the spatial functions of the hippocampus showed that it positions cells involved in head-direction system [44, 45].

Numerous studies have addressed the association of aggressive behaviors with amygdala function. Patients with refractory aggressive behaviors have benefited from amygdala surgeries [46]. Amygdala is considered as the key region for regulating aggressive tendencies [47, 48]. It has been shown that aggressive behaviors are correlated with the size of the amygdala. A negative association was reported between amygdala size and aggressiveness in healthy individuals [49]. On the other hand, several studies have suggested that CD patients have lower amygdala size as well as higher aggressive tendencies. A few studies have attributed the CD-induced aggression to changes in amygdala structure and function [50, 51]. This study reveals that aggressive behaviors in subjects with CC is negatively correlated with the size of amygdala basal nucleus but not with its function.

Several limitations should be considered. First, the limited sample size restricted the generalizability of the results. Second, the cross-sectional setting of this study prevents us from understanding the specific disease course over subjects’ lifetimes. Studying the children with CD in a prospective cohort design could overcome this limitation.

In conclusion, we, for the first time, evaluated the behavioral, emotional, and neurobiological performance of healthy adults with a previous history of CC. Examining the brain structure in this population, we found enlarged left hippocampus and a negative association between volumes of the basal nucleus of left amygdala and the presubiculum of left hippocampus and aggressive behaviors. This study suggests that CC does necessarily progress to antisocial personality disorder; however, some antisocial traits remain in adulthood. Further studies are required to confirm our preliminary results.

## Data Availability

The data used in this project has been obtained from the Human Connectome Project (db.humanconnectome.org). The codes used to analyze the data are available upon request from the corresponding or first author.

https://db.humanconnectome.org

## Acknowledgments

Data were provided by the Human Connectome Project, MGH-USC Consortium (Principal Investigators: Bruce R. Rosen, Arthur W. Toga and Van Wedeen; U01MH093765) funded by the NIH Blueprint Initiative for Neuroscience Research grant; the National Institutes of Health grant P41EB015896; and the Instrumentation Grants S10RR023043, 1S10RR023401, 1S10RR019307.

