## Supplementary Material for "Childhood Conduct History is Linked to Amygdalohippocampal Changes in Healthy Adults: A Neuroimaging Behavioral Study"

**Table 1.** DSM-V criteria for conduct disorder and its corresponding questions from Semi-Structured Assessment for the Genetics of Alcoholism (SSAGA) questionnaire acquired by Human Connectome Project (HCP). We identify subjects with prior history of conduct during their childhood (CC) with at least 3 out of 15 DSM-V conduct symptoms reported present for at least 6 months. “ASP TALLY SHEET PART A” from HCP’s SSAGA was used to identify the presence of the symptoms below for at least 6 months. Note the logical (OR and AND) options applied for each SSAGA questions to fully comply with DSM-V.

|  | **DSM-V Conduct Criteria** | **SSAGA Question No. of HCP** | **SSAGA Question** |
| --- | --- | --- | --- |
| 1 | Often bullies, threatens, or intimidate others | AS9 | Did people complain that you were often a bully, deliberately hurting, threatening, or being mean to other children? |
| 2 | Often initiates physical fights | AS6  OR  AS6B | Did you 3 or more times start physical fights with your brothers or sisters? |
|  |  |  | Did you 3 or more times start physical fights with persons other than your brothers and sisters? |
| 3 | Has used a weapon that can cause serious physical harm to others (e.g., a bat, brick, broken bottle, knife, gun) | AS20 | Did you ever use a weapon like a stick, gun, or a knife to injure someone (other than in combat or as part of your job)? |
| 4 | has been physically cruel to people | AS19 | (Outside of fighting) have you ever physically injured anyone on purpose? |
| 5 | has been physically cruel to animals | AS10 | Did you ever hurt or injure a pet or any other animal on purpose? |
| 6 | has stolen while confronting a victim (e.g., mugging, purse snatching, extortion, armed robbery) | AS16 | Have you ever taken money or property from someone else by threatening them or using force, like snatching a purse or robbing them? |
| 7 | has forced someone into sexual activity | AS21 | Have you ever forced anyone into any sexual activity? |
| 8 | has deliberately engaged in fire setting with the intention of causing serious damage | AS17 | Did you ever deliberately set fires you were not supposed to? |
| 9 | has deliberately destroyed others’ property (other than by fire setting) | AS18 | Have you ever damaged someone's property on purpose  (other than by fire setting)? |
| 10 | has broken into someone else’s house, building, or car | AS15 | Did you ever break into someone else's home, car, or building (not because you were locked out)? |
| 11 | often lies to obtain goods or favors or to avoid obligations (i.e., “cons” others) | AS11a | Did you often lie to get your own way, or to get out of trouble? |
| 12 | has stolen items of nontrivial value without confronting a victim (e.g., shoplifting, but without breaking and entering; forgery) | AS14  OR  AS14b  OR  AS14d | Did you more than once steal money or things from your family, friends, or relatives? COUNT ONLY IF MORE THAN A FEW DOLLARS. |
|  |  |  | Did you more than once steal or shoplift from stores or from other people? (NO CONFRONTATION) |
|  |  |  | Did you more than once forge anyone's signature on a check or use someone’s credit card without permission? |
| 13 | often stays out at night despite parental prohibitions, beginning before age 13 years | AS4  AND  AS_ao4 | Did you ever stay out late at night without permission, either for 2 or more hours after the curfew your parents set or all night without permission? |
|  |  |  | How old were you the first time? <13 |
| 14 | has run away from home overnight at least twice while living in parental or parental surrogate home (or once without returning for a lengthy period) | AS3  AND  (AS3b  OR  AS3C  OR  AS3C1) | Did you ever run away from home overnight? |
|  |  |  | Did you run away overnight more than once? |
|  |  |  | After you ran away, did you return home? |
|  |  |  | When you ran away, how long did you stay away from home? CHECK TALLY IF AWAY FOR 7 OR MORE DAYS |
| 15 | is often truant from school, beginning before age 13 years | AS1  AND  AS3b | Except for your senior year in high school, did you ever play hooky from school for an entire day? |
|  |  |  | How old were you the first time you played hooky twice in one year? MARK TALLY IF AGE ONSET BEFORE 13. |

**Table 2.** Cortical thickness measures shown as millimeters (mean (SD)) based on Desikan-Killiany Atlas. *p-*values are for a general linear model to identify differences between groups, with subjects age and gender as covariates. CC = Childhood Conduct history, HC = No Childhood Conduct History.

| Cortical Parcellation | HC  (n = 1166) | CC  (n = 40) | Uncorrected *p* value | Benjamini-Hochberg Corrected *p* value |
| --- | --- | --- | --- | --- |
| Left banks of superior temporal sulcus | 2.68 (0.14) | 2.68 (0.14) | 0.369 | 0.987 |
| Left caudal anterior cingulate | 2.69 (0.17) | 2.69 (0.20) | 0.552 | 0.960 |
| Left caudal middle frontal | 2.73 (0.12) | 2.75 (0.10) | 0.328 | 0.960 |
| Left cuneus | 2.08 (0.11) | 2.08 (0.11) | 0.749 | 0.960 |
| Left entorhinal | 3.29 (0.24) | 3.31 (0.27) | 0.987 | 0.960 |
| Left frontal pole | 2.84 (0.20) | 2.82 (0.21) | 0.231 | 0.960 |
| Left fusiform | 2.88 (0.12) | 2.87 (0.11) | 0.828 | 0.960 |
| Left inferior parietal | 2.57 (0.11) | 2.58 (0.10) | 0.930 | 0.960 |
| Left inferior temporal | 2.94 (0.13) | 2.95 (0.13) | 0.588 | 0.960 |
| Left Insula | 3.06 (0.15) | 3.07 (0.14) | 0.496 | 0.960 |
| Left isthmus cingulate | 2.28 (0.30) | 2.28 (0.29) | 0.393 | 0.960 |
| Left lateral occipital | 2.29 (0.11) | 2.30 (0.10) | 0.981 | 0.960 |
| Left lateral orbitofrontal | 2.81 (0.12) | 2.81 (0.11) | 0.547 | 0.960 |
| Left lingual | 2.17 (0.11) | 2.17 (0.11) | 0.983 | 0.960 |
| Left medial orbitofrontal | 2.57 (0.15) | 2.57 (0.14) | 0.427 | 0.987 |
| Left middle temporal | 2.98 (0.13) | 2.97 (0.13) | 0.641 | 0.960 |
| Left paracentral | 2.55 (0.13) | 2.54 (0.12) | 0.415 | 0.960 |
| Left parahippocampal | 2.72 (0.26) | 2.69 (0.26) | 0.620 | 0.960 |
| Left parsopercularis | 2.78 (0.12) | 2.79 (0.12) | 0.342 | 0.960 |
| Left pars orbitalis | 2.78 (0.15) | 2.78 (0.12) | 0.138 | 0.960 |
| Left pars triangularis | 2.63 (0.13) | 2.63 (0.10) | 0.051 | 0.987 |
| Left pericalcarine | 2.00 (0.12) | 1.98 (0.13) | 0.940 | 0.960 |
| Left postcentral | 2.21 (0.10) | 2.23 (0.10) | 0.121 | 0.960 |
| Left posterior cingulate | 2.56 (0.13) | 2.54 (0.18) | 0.103 | 0.960 |
| Left precentral | 2.73 (0.11) | 2.76 (0.12) | 0.259 | 0.960 |
| Left precuneus | 2.52 (0.11) | 2.53 (0.10) | 0.959 | 0.960 |
| Left rostral anterior cingulate | 3.03 (0.18) | 3.03 (0.17) | 0.633 | 0.987 |
| Left rostral middle frontal | 2.57 (0.12) | 2.58 (0.10) | 0.065 | 0.960 |
| Left superior frontal | 2.84 (0.13) | 2.84 (0.11) | 0.770 | 0.960 |
| Left superior parietal | 2.29 (0.10) | 2.28 (0.09) | 0.208 | 0.960 |
| Left superior temporal | 2.89 (0.13) | 2.88 (0.12) | 0.565 | 0.960 |
| Left supramarginal | 2.65 (0.11) | 2.66 (0.11) | 0.806 | 0.960 |
| Left temporal pole | 3.42 (0.27) | 3.36 (0.27) | 0.292 | 0.960 |
| Left transverse temporal | 2.66 (0.17) | 2.60 (0.16) | 0.400 | 0.960 |
| Right banks of superior temporal sulcus | 2.79 (0.14) | 2.77 (0.13) | 0.173 | 0.960 |
| Right caudal anterior cingulate | 2.52 (0.23) | 2.53 (0.26) | 0.726 | 0.960 |
| Right caudal middle frontal | 2.75 (0.12) | 2.77 (0.11) | 0.265 | 0.960 |
| Right cuneus | 2.09 (0.12) | 2.08 (0.11) | 0.507 | 0.960 |
| Right entorhinal | 3.42 (0.24) | 3.43 (0.31) | 0.266 | 0.960 |
| Right frontal pole | 2.86 (0.20) | 2.83 (0.20) | 0.079 | 0.960 |
| Right fusiform | 2.90 (0.11) | 2.92 (0.12) | 0.896 | 0.960 |
| Right inferior parietal | 2.65 (0.10) | 2.65 (0.09) | 0.570 | 0.960 |
| Right inferior temporal | 2.97 (0.12) | 3.00 (0.11) | 0.776 | 0.960 |
| Right Insula | 3.01 (0.14) | 3.01 (0.16) | 0.844 | 0.960 |
| Right isthmus cingulate | 2.32 (0.17) | 2.31 (0.17) | 0.567 | 0.960 |
| Right lateral occipital | 2.34 (0.11) | 2.35 (0.12) | 0.538 | 0.960 |
| Right lateral orbitofrontal | 2.83 (0.12) | 2.82 (0.14) | 0.873 | 0.960 |
| Right lingual | 2.19 (0.12) | 2.19 (0.14) | 0.350 | 0.960 |
| Right medial orbitofrontal | 2.74 (0.13) | 2.75 (0.15) | 0.034 | 0.960 |
| Right middle temporal | 3.05 (0.12) | 3.08 (0.12) | 0.998 | 0.960 |
| Right paracentral | 2.58 (0.13) | 2.59 (0.13) | 0.199 | 0.960 |
| Right parahippocampal | 2.70 (0.21) | 2.66 (0.23) | 0.734 | 0.960 |
| Right pars opercularis | 2.82 (0.12) | 2.85 (0.12) | 0.825 | 0.960 |
| Right pars orbitalis | 2.84 (0.15) | 2.82 (0.14) | 0.500 | 0.960 |
| Right pars triangularis | 2.69 (0.12) | 2.74 (0.12) | 0.113 | 0.960 |
| Right pericalcarine | 2.00 (0.12) | 1.99 (0.12) | 0.800 | 0.960 |
| Right postcentral | 2.24 (0.10) | 2.25 (0.12) | 0.119 | 0.960 |
| Right posterior cingulate | 2.53 (0.15) | 2.55 (0.16) | 0.564 | 0.960 |
| Right precentral | 2.73 (0.11) | 2.74 (0.14) | 0.154 | 0.960 |
| Right precuneus | 2.55 (0.11) | 2.54 (0.10) | 0.529 | 0.960 |
| Right rostral anterior cingulate | 3.01 (0.20) | 3.06 (0.18) | 0.877 | 0.960 |
| Right rostral middle frontal | 2.59 (0.11) | 2.60 (0.12) | 0.055 | 0.960 |
| Right superior frontal | 2.87 (0.13) | 2.87 (0.12) | 0.520 | 0.960 |
| Right superior parietal | 2.32 (0.10) | 2.33 (0.09) | 0.343 | 0.960 |
| Right superior temporal | 2.94 (0.13) | 2.97 (0.14) | 0.311 | 0.960 |
| Right supramarginal | 2.70 (0.11) | 2.70 (0.11) | 0.252 | 0.960 |
| Right temporal pole | 3.65 (0.30) | 3.61 (0.35) | 0.630 | 0.960 |
| Right transverse temporal | 2.74 (0.17) | 2.73 (0.17) | 0.717 | 0.960 |

**Table 3.** Volume measures shown as cubic millimeters (mean (SD)) based on Desikan-Killiany Atlas. *p-*values are for a general linear model to identify differences between groups, with subjects age, gender, and total intracranial volume as covariates. CC = Childhood Conduct history, HC = No Childhood Conduct History.

| Volume | HC  (n = 1166) | CC  (n = 40) | Uncorrected *p* value | Benjamini-Hochberg Corrected *p* value |
| --- | --- | --- | --- | --- |
| 3^rd^ Ventricle | 767.62 (242.06) | 715.41 (157.14) | 0.017 | 0.249 |
| 4^th^ Ventricle | 1759.28 (597.71) | 1857.76 (758.40) | 0.647 | 0.964 |
| 5^th^ Ventricle | 4.50 (5.96) | 3.38 (4.58) | 0.210 | 0.860 |
| BrainStem | 21835.24 (2481.91) | 22031.95 (2785.48) | 0.592 | 0.964 |
| Anterior Part of Corpus Callusom | 893.77 (143.04) | 894.97 (161.19) | 0.902 | 0.991 |
| Central Part of Corpus Callusom | 500.58 (107.75) | 544.57 (116.90) | 0.020 | 0.249 |
| Midanterior Part of Corpus Callusom | 495.48 (104.25) | 514.16 (105.97) | 0.316 | 0.964 |
| Midposterior Part of Corpus Callusom | 467.06 (99.27) | 476.70 (104.06) | 0.560 | 0.964 |
| Posterior Part of Corpus Callusom | 963.16 (147.74) | 954.86 (171.07) | 0.746 | 0.991 |
| Cerebrospinal fluid | 1061.66 (216.24) | 1072.73 (193.35) | 0.584 | 0.964 |
| Left Accumbens | 563.96 (93.39) | 577.62 (91.51) | 0.911 | 0.991 |
| Left Amygdala | 1553.94 (205.23) | 1605.97 (173.91) | 0.727 | 0.991 |
| Left Caudate | 3808.37 (475.20) | 3827.19 (513.89) | 0.838 | 0.991 |
| Left Cerebellum Cortex | 57026.10 (6087.67) | 57868.32 (6588.41) | 0.645 | 0.964 |
| Left Cerebellum White Matter | 14562.46 (1892.13) | 14956.08 (2693.87) | 0.646 | 0.964 |
| Left Choroid Plexus | 1123.90 (230.70) | 1077.38 (178.08) | 0.027 | 0.262 |
| Left Hippocampus | 4414.71 (483.37) | 4622.97 (528.47) | 0.032 | 0.262 |
| Left Inferior Horn of Lateral Ventricle | 220.46 (134.16) | 199.62 (124.76) | 0.175 | 0.859 |
| Left Lateral Ventricle | 6521.24 (3647.44) | 5212.46 (2039.11) | 0.005 | 0.247 |
| Left Pallidum | 1363.07 (234.88) | 1321.59 (260.73) | 0.066 | 0.400 |
| Left Putamen | 5522.79 (729.43) | 5413.05 (847.78) | 0.051 | 0.358 |
| Left Thalamus | 8449.50 (918.06) | 8713.73 (868.49) | 0.211 | 0.860 |
| Left Ventral Diencephalon | 4195.33 (490.78) | 4264.16 (656.51) | 0.932 | 0.991 |
| Left Vessels | 65.80 (38.63) | 64.19 (33.31) | 0.485 | 0.964 |
| Left White Mater | 220599.87 (27939.02) | 225097.49 (26991.41) | 0.807 | 0.991 |
| Left Cortical Grey Matter | 252078.21 (26850.87) | 258169.11 (23994.20) | 0.529 | 0.964 |
| Optic Chiasm | 230.68 (53.35) | 241.03 (59.41) | 0.900 | 0.991 |
| Right Accumbens | 598.68 (100.14) | 627.89 (90.22) | 0.266 | 0.964 |
| Right Amygdala | 1635.64 (218.37) | 1671.70 (192.87) | 0.884 | 0.991 |
| Right Caudate | 3927.29 (484.53) | 4005.73 (552.62) | 0.552 | 0.964 |
| Right Cerebellum Cortex | 58593.84 (6331.28) | 59851.49 (7381.08) | 0.996 | 0.996 |
| Right Cerebellum White Matter | 14814.42 (2004.09) | 14893.03 (2258.22) | 0.363 | 0.964 |
| Right Choroid Plexus | 1266.34 (287.95) | 1199.70 (178.51) | 0.011 | 0.249 |
| Right Hippocampus | 4483.10 (457.57) | 4600.89 (510.61) | 0.463 | 0.964 |
| Right Inferior Horn of Lateral Ventricle | 235.27 (149.81) | 231.81 (143.39) | 0.493 | 0.964 |
| Right Lateral Ventricle | 6027.38 (3265.96) | 5310.08 (2452.31) | 0.073 | 0.400 |
| Right Pallidum | 1494.45 (198.89) | 1517.19 (245.89) | 0.942 | 0.991 |
| Right Putamen | 5580.52 (642.31) | 5638.03 (522.98) | 0.582 | 0.964 |
| Right Thalamus | 7389.93 (777.70) | 7557.97 (814.13) | 0.523 | 0.964 |
| Right Ventral Diencephalon | 4232.15 (478.80) | 4303.65 (610.14) | 0.985 | 0.996 |
| Right Vessels | 75.04 (37.00) | 78.14 (38.03) | 0.950 | 0.991 |
| Right White Mater | 223777.16 (28349.49) | 227769.81 (26732.30) | 0.633 | 0.964 |
| Right Cortical Grey Matter | 257539.39 (27451.30) | 263544.46 (24801.82) | 0.628 | 0.964 |
| Subcortical Grey Matter | 60899.11 (5454.32) | 61931.38 (5984.86) | 0.872 | 0.991 |
| SupraTentorial Volume | 1031855.11 (112600.69) | 1051386.86 (103995.78) | 0.836 | 0.991 |
| Total White Matter | 444377.02 (56249.72) | 452867.32 (53676.33) | 0.716 | 0.991 |
| Total Grey Matter | 685480.32 (67142.47) | 700628.76 (62955.15) | 0.649 | 0.964 |
| Total Cortical Grey Matter | 509617.61 (54087.06) | 521713.68 (48719.95) | 0.572 | 0.964 |
| White Matter Hypointensities | 852.77 (411.54) | 848.27 (321.48) | 0.623 | 0.964 |
